# The effects of creatine supplementation on cognitive performance - a randomised controlled study

**DOI:** 10.1101/2023.04.05.23288194

**Authors:** Julia Fabienne Sandkühler, Xenia Kersting, Annika Faust, Eva Kathrin Königs, George Altman, Ulrich Ettinger, Silke Lux, Alexandra Philipsen, Helge Müller, Jan Brauner

## Abstract

**Background:** Creatine is an organic compound that facilitates the recycling of energy-providing adenosine triphosphate (ATP) in muscle and brain tissue. It is a safe, well-studied supplement for strength training. Previous studies have shown that supplementation increases brain creatine levels, which might increase cognitive performance. The results of studies that have tested cognitive performance differ greatly, possibly due to different populations, supplementation regimens and cognitive tasks. This is the largest study on the effect of creatine supplementation on cognitive performance to date. As part of our study, we replicated Rae et al. (2003).

**Methods:** Our trial was cross-over, double-blind, placebo-controlled, and randomised, with daily supplementation of 5g for six weeks each. Like Rae et al. (2003), we tested participants on Raven’s Advanced Progressive Matrices (RAPM) and on the Backward Digit Span (BDS). In addition, we included eight exploratory cognitive tests. About half of our 123 participants were vegetarians and half were omnivores.

**Results:** There was no indication that vegetarians benefited more from creatine than omnivores, so we merged the two groups. Participants’ scores after creatine and after placebo differed to an extent that was not statistically significant (BDS: p = 0.064, η^2^_P_ = 0.029; RAPM: p = 0.327, η^2^_P_ = 0.008). Compared to the null hypothesis of no effect, Bayes factors indicate weak evidence in favour of a small beneficial creatine effect and strong evidence against a large creatine effect. There was no indication that creatine improved the performance of our exploratory cognitive tasks. Side effects were reported significantly more often for creatine than for placebo supplementation (p = 0.002, RR = 4.25).

**Conclusions:** Our results do not support large effects of creatine on the selected measures of cognition. However, our study, in combination with the literature, implies that creatine might have a small beneficial effect. Larger studies are needed to confirm or rule out this effect. Given the safety and broad availability of creatine, this is well worth investigating; a small effect could have large benefits when scaled over time and over many people.

## Introduction

Given the important role cognition plays in daily life, substances that enhance cognition safely and cheaply are highly desirable. Creatine is safe, well-tolerated, and cheap (Kreider et al., 2017). Strength athletes have benefited from creatine supplementation for over 30 years (Branch, 2003; Butts et al., 2018). Slight weight gain due to water retention is the only consistently reported side effect (Bender et al., 2008; de Souza e Silva et al., 2019; Kreider et al., 2017; Kutz & Gunter, 2003).

While the safety and athletic benefits of creatine are well established, its potential cognitive benefits are still unclear. A systematic review tentatively suggests that creatine supplementation may improve “short-term memory”/working memory and “intelligence/reasoning” in healthy individuals (Avgerinos et al., 2018). The few studies that have tested this have had heterogeneous results, but they have also used very different populations (such as vegetarians, omnivores, varying age groups), supplementation doses and durations and cognitive tasks (including different kinds of memory, reaction time, reasoning, inhibitory control, attention and task switching). The study with the largest effect, Rae et al. (2003), tested the effect of creatine supplementation in 45 young vegetarian adults on working memory and abstract reasoning using the Backwards Digit Span (BDS) and Raven’s Advanced Progressive Matrices (RAPM), respectively. Their study was placebo-controlled, randomised and double-blind. Rae et al. (2003) found creatine supplementation had a large and highly significant positive effect on both tasks. We deemed this study particularly worth replicating.

Why might supplementing creatine benefit cognition? Muscle and brain cells use creatine to access more energy when demand is high. They store creatine as phosphocreatine, which acts to regenerate the energy-providing adenosine triphosphate (ATP) (Lowe et al., 2013; Persky & Brazeau, 2001). The energy demand of neurons can increase rapidly; maintaining ATP concentration despite increased demand may explain the potential effect of creatine intake on cognition (Ainsley Dean et al., 2017). The crucial role of creatine in brain metabolism is supported by evidence from Cerebral Creatine Deficiency Syndromes. Conditions causing brain creatine deficiency result in profound intellectual disability which can be reversed by creatine supplementation (Clark & Cecil, 2015).

Creatine can be produced by the body and is present in common foods, so why would we expect supplementation to make a difference? Dietary creatine is primarily contained in meat, fish, and a small amount in some dairy products (Balestrino & Adriano, 2019; Brosnan & Brosnan, 2016). However, typical supplementation doses of creatine (5g per day) are equivalent to more than 1kg of meat consumption per day (Brosnan & Brosnan, 2016), which is substantially higher than the combined dietary intake and synthesis in most people (Brosnan & Brosnan, 2016).

Creatine intake increases the level of creatine in the blood serum (Harris et al., 1992) (Schedel et al., 1999). Crucially, (Dechent et al., 1999) found brain creatine increased by 8.7% following a 20g/day 4-week supplementation regime; two further studies have confirmed varying supplementation regimes can increase brain creatine (Lyoo et al., 2003; Turner, Russell, et al., 2015).

It is unclear if creatine supplementation has similar effects on omnivores and vegetarians. Rae et al. (2003) only included vegetarians. Another study comparing memory improvement under creatine supplementation in omnivores and vegetarians found that creatine supplementation improved memory only for vegetarians but not omnivores (Benton & Donohoe, 2011). Vegetarians have been found to have lower serum and muscle creatine concentration, but comparable total brain creatine to omnivores (Burke et al., 2003; Solis et al., 2014, 2017). In this study, we included both omnivores and vegetarians to allow comparison. We hypothesised that creatine supplementation would improve working memory and reasoning ability in vegetarians. We also hypothesised that the improvement would be greater in vegetarians than in omnivores.

To test these hypotheses, we approximately replicated the study design and treatment (5g per day of creatine for six weeks) used by Rae et al. (2003). We included the same primary outcome measures, the Backwards Digit Span and 10-minute standardised subtests of Raven’s Advanced Progressive Matrices. In addition, to investigate a broader range of cognitive functions, we included exploratory tests on attention, verbal fluency, task switching, and memory.

## Methods

### Trial design

We conducted a randomised, placebo-controlled, double-blind, cross-over study. The primary endpoints are the scores in the cognitive tasks after 6 weeks of each supplementation. Six weeks have been found to be a sufficient wash-out period (private correspondence, (Turner, Byblow, et al., 2015)) and it is the duration used by Rae et al. (2003). Unlike Rae et al. (2003), we did not have an extra washout period nor second baseline testing after the first supplementation. The trial evaluated cognitive performance after creatine compared to placebo. The trial design and participant flow are summarised in Figure 1. The trial was prospectively registered (drks.de identifier: DRKS00017250, https://osf.io/xpwkc/) and ethical approval was obtained from the ethics committee of the University of Bonn (060/19). We follow the CONSORT guidelines.

**Figure 1.**
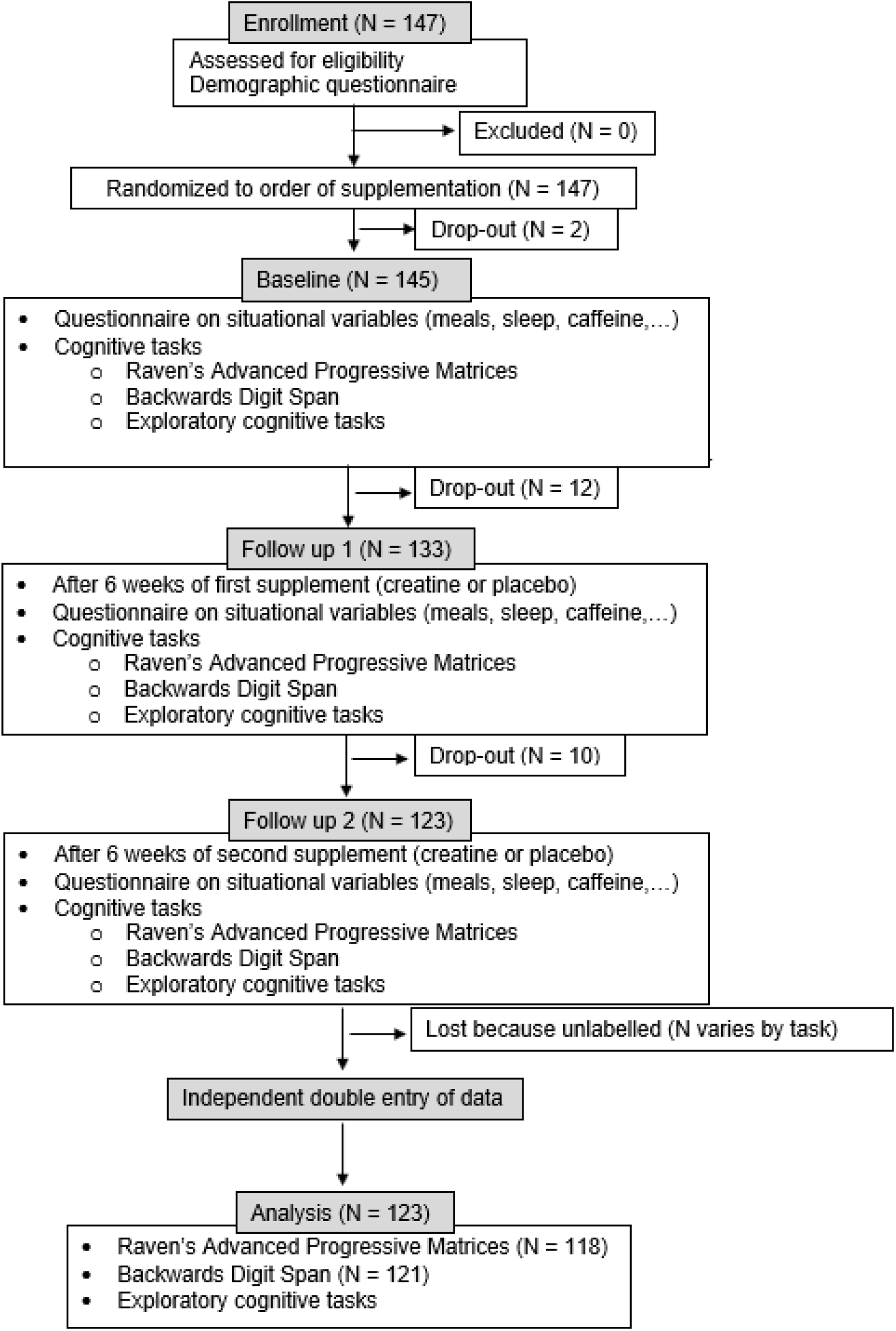
Participant flow through the study.

### Participants

Participants were 18 years or older (see appendix for full list of inclusion criteria). Half of them reported to be on a vegetarian diet and half of them on an omnivore diet. Cognitive assessments of participants took place in the clinic laboratory. Due to the contact restrictions due to the COVID-19 pandemic, after 04/2020 participants were tested online via video call instead. A screening questionnaire assessed if the eligibility criteria were met. Participants who met these criteria went through the baseline assessment and were given their first supplement to take home (or for participants tested online, received the two supplements via the mail).

### Interventions and similarity of treatment groups

Participants took the supplements daily for six weeks, including the day of the testing. The creatine supplement consisted of creatine monohydrate powder “CreaPure PG” produced by the company Alzchem (Trostberg, Germany). The placebo supplement consisted of maltodextrin powder “Maltodextrin 6” produced by the company Nutricia (Frankfurt am Main, Germany.

The cans looked exactly the same except for clear markings of which one was the first and which one the second supplement. The two powders looked exactly the same and were flavourless. The solubility was somewhat different: While the placebo powder was completely soluble in water and did not settle, the creatine powder slowly settled. We were initially not aware of this difference in solubility. After we noticed it (after the first 40 participants), we asked participants to stir the powder into yoghurt or food with a similar consistency, as we had found no perceptible difference then. To check to what extent blinding was achieved, directly after the last testing participants were asked to guess what their first supplement had been.

### Outcomes

We had two primary outcomes:

- A standardised 10-minute subtest of Raven Advanced Progressive Matrices (RAPM) (Rae et al., 2003)
- The Wechsler auditory Backward Digit Span (BDS) (Wechsler 1955)

RAPM is a test of abstract reasoning. Each item in the test consists of a 3×3 matrix with pictures of geometric forms. One of the pictures is missing and the task consists of choosing the right picture to fill this gap out of eight alternatives. The full RAPM consists of 80 items and has a time limit of 40 minutes. We used the same standardised 10-minute subtests of the RAPM as Rae et al. (2003), consisting of 20 items each. The subtests are constructed to have equal levels of difficulty based on the published normative performance data and Rae et al. (2003) additionally verified this in an independent sample (N = 20). The RAPM score consists of the sum of correct responses.

The Backward Digit Span is a test of working memory. The tester reads increasingly longer series of digits to the participant whose task it is to remember and repeat them in reverse order. The task starts with two digits. Each length has two series of digits. The test ends after wrong answers to two series of the same length. The BDS score consists of the sum of correct responses.

We had eight further exploratory outcomes:

- The D2 Test of Attention (Brickenkamp, 2002), a test of sustained attention
- The Trail-Making-Test A (TMT-A), a test of visual attention (Reitan, 1958)
- The Trail-Making-Test B (TMT-B), a test of task switching (Reitan, 1958)
- The Block-Tapping-Test (BTT), a test of visuospatial working memory (Schellig, 1997)
- The Auditory Verbal Learning Test (AVLT, in German: VLMT), a word-learning test including immediate recall, delayed recall and recognition (Lux et al., 2001)
- The Brief-Visuospatial-Memory Test – Revised (BVMT-R), a test of visuospatial memory (Benedict et al., 1996)
- The Stroop test (in German: Farb-Wort-Interferenz Test, FWI-T), a test of inhibitiory control (Bäumler & Stroop, 1985)
- Regensburger Wortflüssigkeitstest (RWT), a test of verbal fluency (Aschenbrenner et al., 2000)

Participants reported side effects experienced during the supplementation period in a free text form on the day of testing. How side effects would be grouped for the report was determined after evaluating all entries. At baseline testing, participants performed a test of crystallised intelligence called “Mehrfach-Wahl-Wortschatztest (MWT-B)” (Lehrl, 2005). In this test, participants had to identify real German words among made-up words.

### Sample size

The sample size of 123 was powered (with power = 0.8, alpha = 0.05, calculated with GPower) to detect effects of Cohen’s d = 0.45. The sample size (preregistered as 120) was chosen based on a conservative estimate (see appendix) of the effect size in Rae et al. (2003) (d = 1 for both RAPM and BDS) with a substantial buffer to account for smaller effects.

Block-tapping was originally performed with physical blocks and later on the website Psytoolkit (Stoet, 2010, 2017) as part of remote testing during the COVID-19 pandemic. Because the remote version was not immediately available, the participant number is lower for this task.

### Randomisation and blinding

The order of the two supplements was randomised with Excel by the pharmacy of the university hospital Heidelberg. They labelled each of the cans of supplements with the participant code and “A” or “B”, corresponding to the first and second supplement. The staff members who tested participants also provided the participants with the supplement cans. Allocation concealment was performed using sequentially numbered, opaque sealed envelopes (SNOSE). Participants and all staff who interacted with them were kept blinded to the allocation (also see intervention section).

### Statistical methods

For each cognitive test, we conducted a mixed ANOVA with test score after supplementation as the dependent variable, supplement (creatine vs placebo) as the within-subjects factor and supplement order (creatine-first vs placebo-first) as the between-subjects factor. We did not remove outliers in our main analysis, but conducted robustness checks which included trimming and winsorising. We applied the Greenhouse-Geisser correction to all our analyses but the correction did not change any value.

#### Confirmatory analyses

As preregistered, our two confirmatory cognitive tasks are the Backward Digit Span and Raven’s Advanced Progressive Matrices. All other cognitive tasks are analysed in an exploratory fashion.

There is one deviation from our preregistered analyses. We had preregistered t-tests, but this was a mistake in the preregistration. The t-test is not appropriate here because imbalances in the supplement order group sizes would bias the results. Instead we conducted mixed ANOVAs with supplement (creatine vs placebo) as the within-subjects variable, supplement order (creatine-first vs placebo-first) and diet (vegetarian vs omnivore) as the between-subjects variables and test score after supplementation as the dependent variable.

#### Robustness checks

We checked the robustness of our normality-assuming ANOVAs by performing: an ANOVA on 20%-trimmed data, an ANOVA on 5%- and on 20%-winsorised data, and a robust ANOVA which uses trimming and bootstrapping (performed with the sppb functions in the WRS2 R package). The latter ANOVA provides the most robust estimate of these methods (Field, 2013; Wilcox, 2011).

#### Bayes factors

For the calculation of the Bayes factors, we used the estimated marginal means (EMMs) of the creatine and placebo score. The EMMs are the means weighed for the order groups (creatine-first and placebo-first), so that imbalances in the sizes of the order groups do not affect the means. So, we only had two groups for the Bayes factor calculation (creatine and placebo), simplifying the analysis. The mean difference and standard error of the mean difference were used to describe the data. Using the Bayesplay package (Colling, 2021), we calculated the Bayes factors in several different ways. Approach 1 used point models for the null hypothesis and the alternative hypotheses. Approach 2 compared a point null model against half normal distributions centred on zero and with the standard deviation set to half the maximum expected effect size. For the reasons behind this see the appendix.

#### Exploratory analyses

In addition to the confirmatory analyses of BDS and RAPM, we analysed the other cognitive tasks in the same way in an exploratory fashion.

We also looked in an exploratory fashion at the first supplementation and the second supplementation separately and at participants with a low and high baseline performance separately (see appendix).

## Results

### Participant flow

### Recruitment

Participants were recruited through flyers and social media between 05/2019 and 05/2022 and tested between 05/2019 and 08/2022.

### Baseline data

We analysed all available participant data apart from one task in the case of two participants (see appendix). Participants were included irrespective of their adherence. For participant characteristics, see Table 1.

**Table 1.** Participant characteristics. Data is given as mean (standard deviation) or as percentage. The MWT-B (Mehrfach-Wahl-Wortschatztest) is a test of crystallised intelligence (Lehrl, 2005).

|  | Total | Creatine-first | Placebo-first |
| --- | --- | --- | --- |
| Age in years (M, SD) | 30.6 (10.1) | 31.5 (10.4) | 29.8 (9.7) |
| Sex (% female) | 57% | 54% | 60% |
| Weight in kg (M, SD) | 70.3 (13.7) | 71.8 (15.5) | 68.8 (11.4) |
| MWT-B (M, SD) | 26.31 (4.35) | 26.32 (3.99) | 26.31 (4.72) |

### Blinding, adherence and side effects

The last 73 participants were asked to guess the order of their supplements. Forty-three (59%) guessed correctly and 30 (41%) guessed incorrectly. A binomial test reveals that the probability of 43 or more correct guesses out of 73 by pure chance is p = 0.080. However, most participants who guessed correctly reported being very unsure about their guess. We recorded the reasons for the guesses of the last of the 33 participants. Of those participants who had a reason for their guess, solubility was the most common, followed by negative side effects and positive side effects. All three reasons seemed to improve guess accuracy (see appendix).

A z-score test for two population proportions revealed that the proportion of participants reporting any negative side effect was significantly higher for the creatine than the placebo condition, p = 0.002, RR = 4.25 (Table 2). In addition, although we did not assess this systematically, some participants reported positive side effects such as improvements in strength (several participants) and mood (one participant). No patients discontinued the study due to an adverse event.

**Table 2.**
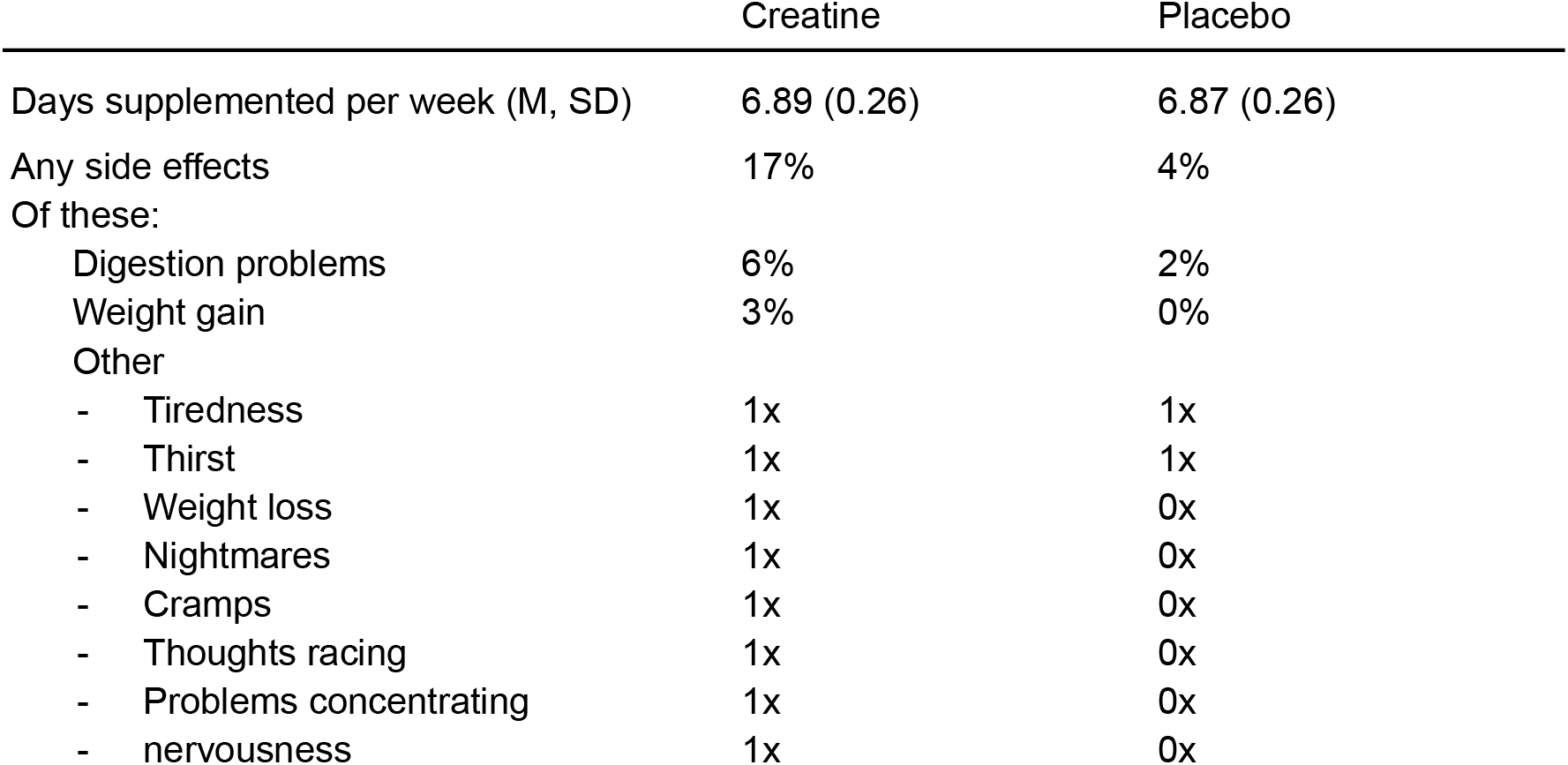
Adherence and negative side effects.

|  | Creatine | Placebo |
| --- | --- | --- |
| Days supplemented per week (M, SD) | 6.89 (0.26) | 6.87 (0.26) |
| Any side effects | 17% | 4% |
| Of these: |  |  |
| Digestion problems | 6% | 2% |
| Weight gain | 3% | 0% |
| Other |  |  |
| - Tiredness | 1x | 1x |
| - Thirst | 1x | 1x |
| - Weight loss | 1x | 0x |
| - Nightmares | 1x | 0x |
| - Cramps | 1x | 0x |
| - Thoughts racing | 1x | 0x |
| - Problems concentrating | 1x | 0x |
| - nervousness | 1x | 0x |

Adherence (self-reported) was high (Table 2). All but one participant took the supplements in the order assigned to them. This participant was analysed with their actual, not their assigned, supplement order.

### Interaction with diet

There was no significant interaction between diet and supplement nor between diet, supplement and supplement order for neither BDS (p = 0.808 and p = 0.559) nor RAPM (p = 0.392 and p = 0.606), nor was the interaction in the predicted direction (we had hypothesised that vegetarian participants would benefit more from creatine than omnivore participants). This was also true when using the robust ANOVA based on bootstrapping. Bayes factors favoured the null hypothesis. To be precise: They indicated strong support in favour of the null hypothesis over the effect size in Benton and Donohoe (2011) (d = 0.36) and weak to strong support in favour of the null hypothesis over smaller effect sizes (see appendix). There was no indication for an effect of diet in the exploratory cognitive tasks either. For more details on the analysis of diet, see the appendix.

### Confirmatory analysis

There was a significant interaction between supplement and supplement order for both BDS and RAPM. This seems to reflect a learning effect (see appendix). The effect of most interest, the main effect of the supplement, was in the expected direction but not significant. However, it bordered on significance for BDS (p = 0.067, η^2^_P_ = 0.028). This means that 2.8% of the variance in BDS scores that was not already explained by other variables was explained by the supplement. For RAPM, it was 0.9%. The supplement effect was virtually the same whether diet was included as a variable or not (Table 3). Thus we simplified additional analyses (estimated marginal means, Bayes factors and robustness checks) by dropping diet as a variable for these analyses.

**Table 3.**
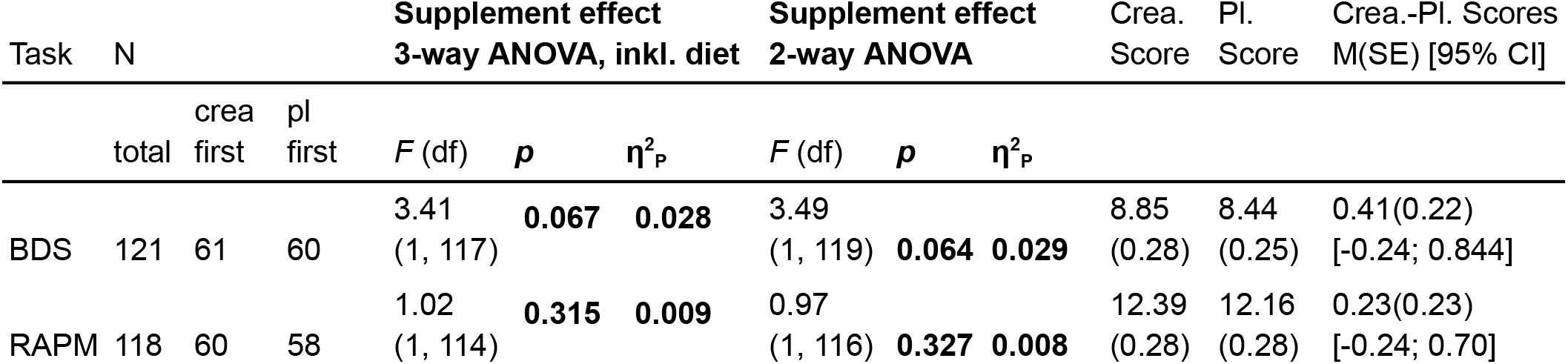
Mixed 3-way ANOVA with supplement (creatine vs placebo) as the within-subjects variable, supplement order (creatine-first vs placebo-first) and diet (vegetarian vs omnivore) as the between-subjects variable and test score after supplementation as the dependent variable. Mixed 2-way ANOVA without diet. The test score is given as estimated marginal mean (standard error). P-values are two-tailed. The two cognitive tasks are the Backward Digit Span and Raven’s Advanced Progressive Matrices.

In terms of raw scores, the effect size for BDS was 0.41 additional correct items, i.e. a 0.2 digits longer digit span, because there were always two digit spans of the same length. For RAPM, the effect was 0.23 more matrices solved (Figure 2). Cohen’s d based on the estimated marginal means of the creatine and placebo scores was 0.09 for RAPM and 0.17 for BDS. If these were IQ tests, this would mean 1.4 and 2.6 IQ points.

**Figure 2.**
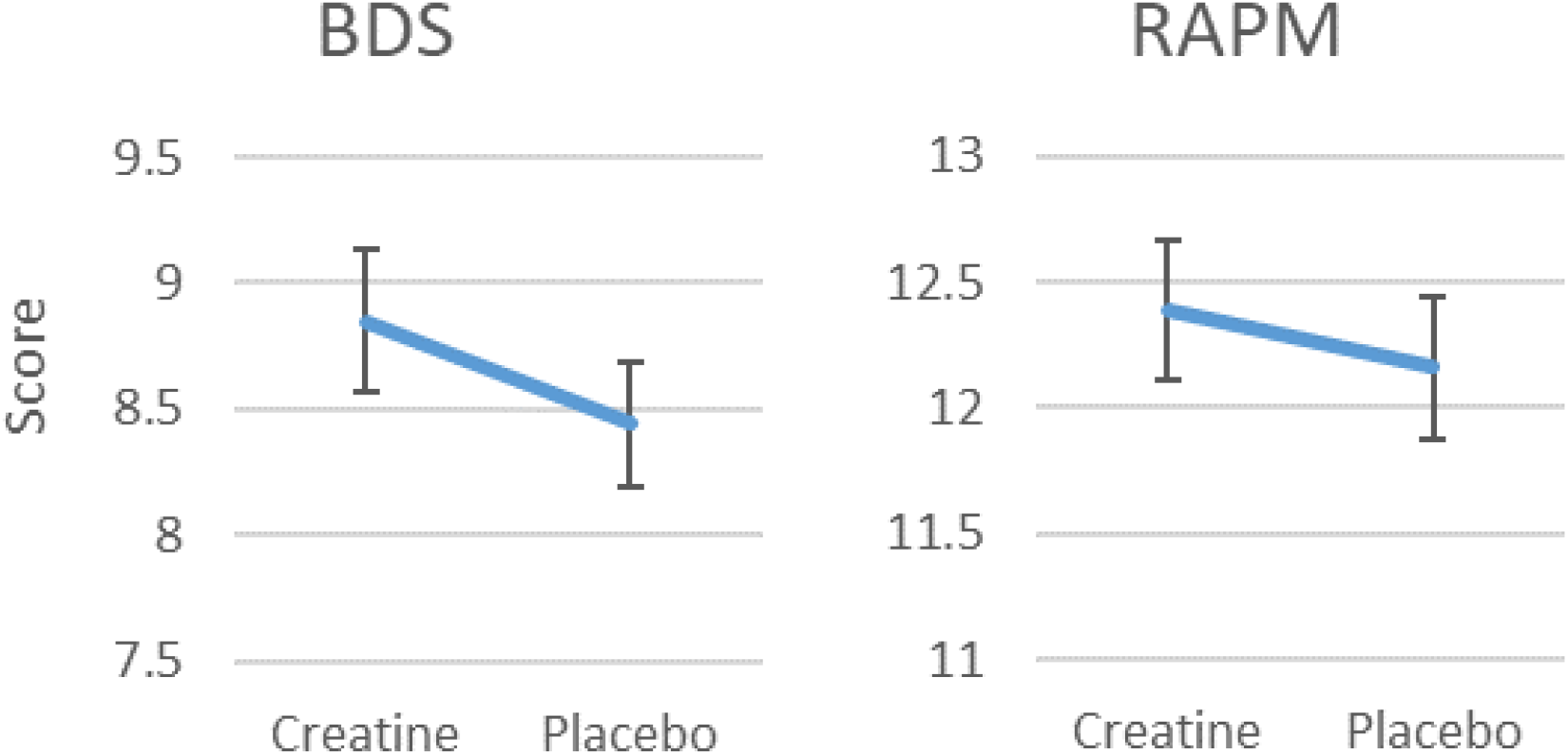
a) Estimated marginal means for the Backward Digit Span (BDS) score. b) Estimated marginal means for Raven’s Advanced Progressive Matrices (RAPM) score. Error bars represent standard errors.

### Bayes factors

To facilitate the interpretation of the results of the confirmatory analysis, we provide Bayes factors. A Bayes factor (BF_10_) indicates how likely a null hypothesis is compared to an alternative hypothesis given the data. A BF_10_ between ⅓ and 3 indicates low sensitivity of the data (i.e. not enough data to be certain), with weak evidence in favour of the null hypothesis if BF_10_ is below 1 and weak evidence in favour of the alternative hypothesis if it is above 1. A BF_10_ above 3 (below ⅓) is considered moderate and above 10 (below 1/10) strong evidence (van Doorn et al., 2021).

We compare several alternative hypotheses postulating small beneficial effects of creatine to the null hypothesis. For RAPM, the data was very insensitive, very weakly favouring the alternative hypotheses. For BDS, the data was more sensitive, providing weak to moderate support in favour of the alternative hypotheses. Two different approaches to calculating these Bayes factors were used (see statistical analysis) and the results were similar (Table 4).

**Table 4.**
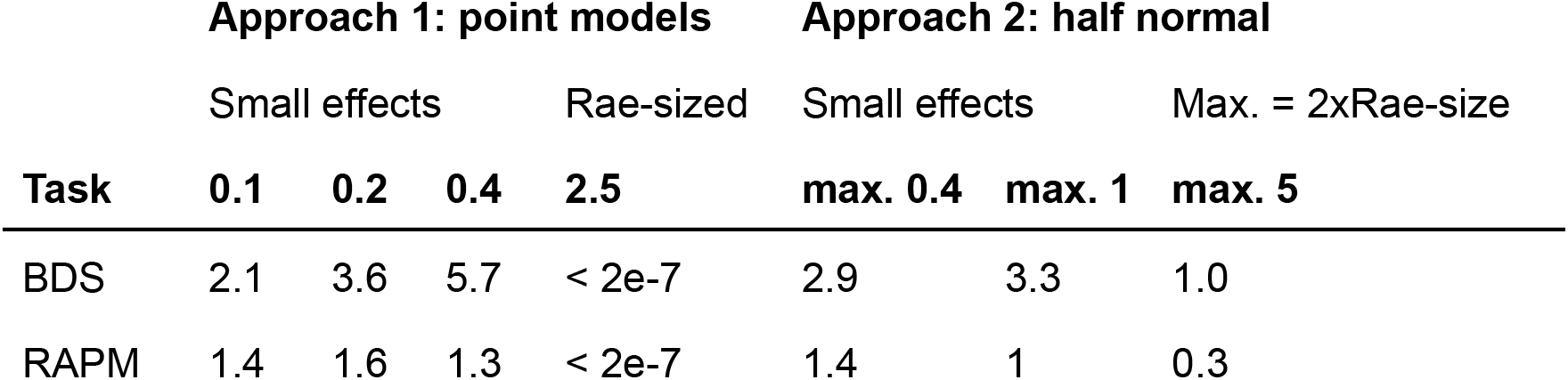
Bayes factors (BF_10_) comparing a range of alternative hypotheses to the null hypothesis. The effect size is given as the raw score difference. Approach 1 compared a point null model to point alternative models with a range of small effect sizes (0.1-0.4, i.e. d = 0.04-0.17) as well as an equivalent of Rae et al.’s effect size (2.5, i.e. d = 1, see calculation in appendix). Approach 2 compared a point null model against half normal distributions centred on zero and with the SD set to half the maximum expected effect size.

There was strong evidence in favour of the null hypothesis compared to the alternative hypothesis postulating the effect size found by Rae et al. (2003). The data was insensitive (BDS) or weakly favoured the null hypothesis (RAPM) when compared to the half normal model based on Rae et al. (2003). The half normal model based on Rae et al. (2003) does not assume their effect size is the true effect size in the population. Instead, the model assumes their effect size is a moderate overestimation of the true effect size. The model uses their effect size as a reference point to assign probabilities to effect sizes. It assigns most of the probability weight to effect sizes that are smaller than this effect size, and some probability to effect sizes up to twice that effect size. This is a common alternative model when replicating studies. However, we did not use it as our only model, because we were also interested in assessing the likelihood of smaller effect sizes and of the possibility that the effect size in Rae et al. (2003) was the true population effect size.

The results were similar whether using normal or cauchy distributions. For more details on this and the aforementioned calculations see the appendix.

In summary, this study provides weak evidence for a small cognitive benefit of creatine and strong evidence against the effect size by Rae et al. (2003) being representative.

### Robustness checks

We checked the robustness of our confirmatory analysis (the normal ANOVA) by performing: an ANOVA on 20%-trimmed data, an ANOVA on 5%- and on 20%-winsorised data, and an ANOVA which uses bootstrapping and 20% trimming.

For RAPM, all of these methods gave overall similar results to that of the normal ANOVA (Table 5).

**Table 5.**
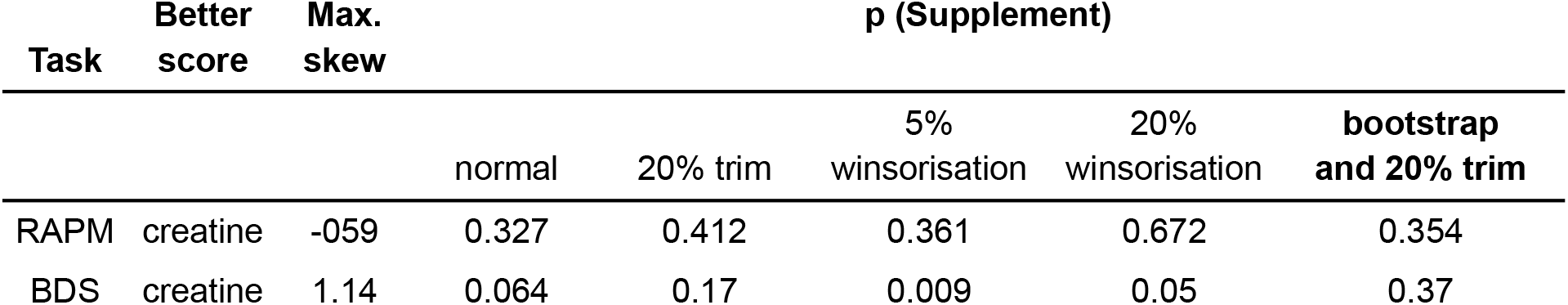
Creatine effect p-values (two-tailed) for different ANOVAs. The given trim and winsorisation percentages are applied to each side. Better score based on estimated marginal means. “Max. skew” gives the highest skewness statistic in any combination of conditions (supplement and supplement order).

| Task | Better score | Max. skew | p (Supplement) |  |  |  |  |
| --- | --- | --- | --- | --- | --- | --- | --- |
|  |  |  | normal | 20% trim | 5% winsorisation | 20% winsorisation | bootstrap and 20% trim |
| RAPM | creatine | -059 | 0.327 | 0.412 | 0.361 | 0.672 | 0.354 |
| BDS | creatine | 1.14 | 0.064 | 0.17 | 0.009 | 0.05 | 0.37 |

For BDS, whose skewness statistic was slightly further from 0 than that of RAPM, these methods gave results that differ from each other and from the normal ANOVA to a relevant extent (Table 5). Most notably, the p-value for the supplement effect was 0.009 for the 5%-winsorisation and 0.370 for the bootstrap ANOVA. This seems to suggest that in the normal ANOVA, the most extreme values made the effect of creatine appear smaller by inflating the variance, while relying on possibly unjustified assumptions of normality made the effect of creatine appear larger.

Thus, the result for RAPM was robust and for BDS much less so.

### Exploratory cognitive tasks

There was no indication that creatine improved the performance of our exploratory cognitive tasks. The distribution of p-values was what one would expect if there was no effect. For the exploratory cognitive tasks, Table 6 only includes the p-values of the supplement effect. For the full results, including the interaction effect (reflecting a learning effect) and the order of supplement effect, see the appendix.

**Table 6.** Creatine effect p-values (two-tailed) for different ANOVAs. The given trim and winsorisation percentages are applied to each side. Higher score based on estimated marginal means.

| Task | N | Better score | p (Supplement) |  |  |  |  |
| --- | --- | --- | --- | --- | --- | --- | --- |
|  |  |  | normal | 20% trim | 5% winsorisation | 20% winsorisation | bootstrap and 20% trim |
| Blocktapping forward | 71 | creatine | 0.779 | 0.564 | 0.865 | 0.678 | 0.826 |
| Blocktapping backward | 70 | placebo | 0.83 | 0.87 | 0.482 | 0.482 | 0.59 |
| BVMT-R | 119 | creatine | 0.543 | 0.809 | 0.746 | 0.112 | 0.67 |
| D2 test | 104 | placebo | 0.394 | 0.382 | 0.446 | 0.291 | 0.62 |
| Forward digit span | 117 | placebo | 0.714 | 0.795 | 0.838 | 0.721 | 0.52 |
| Stroop - colors | 118 | placebo | 0.813 | 0.184 | 0.432 | 0.054 | 0.568 |
| Stroop - colorletters | 119 | placebo | 0.626 | 0.877 | 0.861 | 0.547 | 0.856 |
| TMT A | 123 | placebo | 0.129 | 0.04 | 0.068 | 0.021 | 0.224 |
| TMT B | 122 | creatine | 0.855 | 0.622 | 0.745 | 0.567 | 0.56 |
| VLMT immediate recall | 119 | creatine | 0.87 | 0.744 | 0.996 | 0.883 | 0.996 |
| VLMT recall after interference | 119 | creatine | 0.694 | 0.622 | 0.854 | 0.323 | 0.806 |
| VLMT delayed recall | 118 | placebo | 0.339 | 0.346 | 0.462 | 0.133 | 0.54 |
| VLMT recognition | 117 | creatine | 0.722 | 0.327 | 0.398 | 0.635 | 0.348 |
| Word fluency | 122 | creatine | 0.227 | 0.631 | 0.272 | 0.119 | 0.692 |

## Discussion

This is the largest study on the cognitive effects of creatine to date. As part of our study, we aimed to replicate Rae et al. (2003), who found a large positive effect of creatine on the abstract reasoning task Raven’s Advanced Progressive Matrices (RAPM) and on the working memory task Backward Digit Span in healthy young adult vegetarians.

In our study, half of the participants were vegetarians and half of them were omnivores. We found no indication that our vegetarian participants benefited more from creatine than our omnivore participants. This is in line with Solis et al (2014, 2017) who did not find a difference in brain creatine content between omnivores and vegetarians. Our Bayesian analysis of their data provides moderate support for the lack of a difference (see appendix). In contrast, Benton and Donohoe (2011) found that creatine supplementation benefited memory in vegetarians more than in omnivores, with no difference in baseline performance. However, given the high number of tests in that study, the chance of a false positive was high, so we regard their finding as only an exploratory hint. The conflicting findings might be due to possible differences in the amount of dietary creatine (not measured in this study nor in Benton and Donohoe (2011)).

The preregistered frequentist analysis of RAPM and BDS found no significant effect at p < .05 (two-tailed), although the effect bordered significance for BDS. The Bayes factors in this study provide weak evidence for a small cognitive benefit of creatine and strong evidence against the large effect size found by Rae et al. (2003). A larger sample size would be necessary to provide stronger evidence on the question of a small benefit. In order for the sample size to not have to be exceedingly large, we recommend being extremely careful in reducing noise and choosing participants who are likely to benefit the most. In addition, analogous to the compounding effect of creatine over time for strength training, it might be possible to see a larger effect of creatine on cognition over time by training the tasks while on the supplement.

In their review, Avgerinos et al. (2018) reported that a creatine effect is more likely for “intelligence/reasoning” and “short term memory”/working memory than for other cognitive domains. In line with this, we found a weak indication for a creatine effect for the two confirmatory tasks reflecting these two domains but not for other domains. However, two of our exploratory tasks, the forward digit span and the immediate recall part of the VLMT also tested short-term memory and there was no indication for an effect for these tasks. Another review, (Dolan et al., 2018) report that a creatine effect is more likely for more cognitively demanding tasks. In line with this, we found some indication for a creatine effect for the backward digit span (BDS) but not for the less demanding forwards digit span. The VLMT may also be less cognitively demanding than the BDS, but this comparison is less obvious to make.

There are a number of limitations to this study. Despite the large sample size compared to other studies, a larger sample size would be needed to be powered for effects that are smaller but still relevant. Some of the data (2%) could not be analysed because it was not labelled with the participant and timepoint. The COVID-19 pandemic started in the middle of the study, which likely added noise to the data, and meant that we had to switch from in-person cognitive testing to testing via video call. Adherence was self-reported and not checked with blood samples. Another limitation is that the proportion of participants who correctly guessed their supplement order (59%) bordered on significance (p = 0.08).

However, most participants who guessed correctly reported being very unsure about their guess. The largest contributing factor to correct guesses was likely the difference in the solubility between the powders, followed by negative and positive side effects. We attempted to counteract differences in solubility by recommending participants to stir the supplements into yoghurt. For future studies we recommend cellulose as the placebo and a mixture of cellulose and creatine as the treatment, as these two look extremely similar when dissolved in water. The alternative solution with capsules would require participants to consume many capsules per day. This would likely reduce adherence and massively increase costs.

Unfortunately, it is difficult to achieve perfect blinding when side effects occur with higher frequency in the creatine condition. The side effects of creatine are well-known and not dangerous (Bender et al., 2008; de Souza e Silva et al., 2019; Kreider et al., 2017; Kutz & Gunter, 2003).

## Conclusion

Supplementing creatine is safe, easy and very cheap. The real effect of creatine on cognition is likely smaller than that reported in Rae et al. (2003). However, even small improvements in cognition may be relevant, especially if accumulated over many people and over time. The results of this study do not allow any strong conclusions, but it would be worthwhile to test for a small effect of creatine in strategically designed, larger studies.

## Data Availability

The appendix, data, code and output of this study are openly available at the Open Science Framework, https://osf.io/xpwkc/.

## Acknowledgements

We thank the doctors of the University Clinic Bonn who collected blood samples. We thank Dr. Lincoln Colling, Dr. Christian Stark, Jan Speller, Maximilian Meier and David Reinstein for their feedback on statistical questions. We thank Thomas Szpejewski and Tom Lieberum for their help with verifying data quality. We thank all data entry helpers.

## Funding

Funding was provided by the non-profit organization Effective Ventures Foundation, 2443 Fillmore St., #380-16662, San Francisco, CA 94115. The trial funders had no role in the design of the study, the collection, analysis or interpretation of data, the writing of the report, or the decision to submit the article for publication.

## Notes

### Competing Interest Statement

The authors have declared no competing interest.

### Clinical Trial

DRKS00017250

### Author Declarations

Ethical approval was obtained from the ethics committee of the University of Bonn (060/19).

