## Appendix for "The effects of creatine supplementation on cognitive performance - a randomised controlled study"

#### Inclusion criteria

All inclusion criteria were self-reported.

- No psychological or physical disorders or disabilities if they are likely to cause instabilities in test scores or interact with creatine
- Age of 18 and above
- Stable eating behaviour, i.e., being either omnivore or vegetarian/vegan for at least 6 months
- No creatine intake in the last 6 months before the study begins
- Ability to consent
- Alcohol consumption on average not higher than 20g of alcohol per day (if female) or not more than 40g of alcohol per day (if male)
- No consumption of recreational drugs more than once a week
- Not more than 6 hours of intense sports per week

#### Excluded data

We excluded the data of one Stroop test of one participant due to a technical error.

We excluded the data of one forward and backward digit span test of one participant because they had evidently cheated.

### Histograms of BDS scores

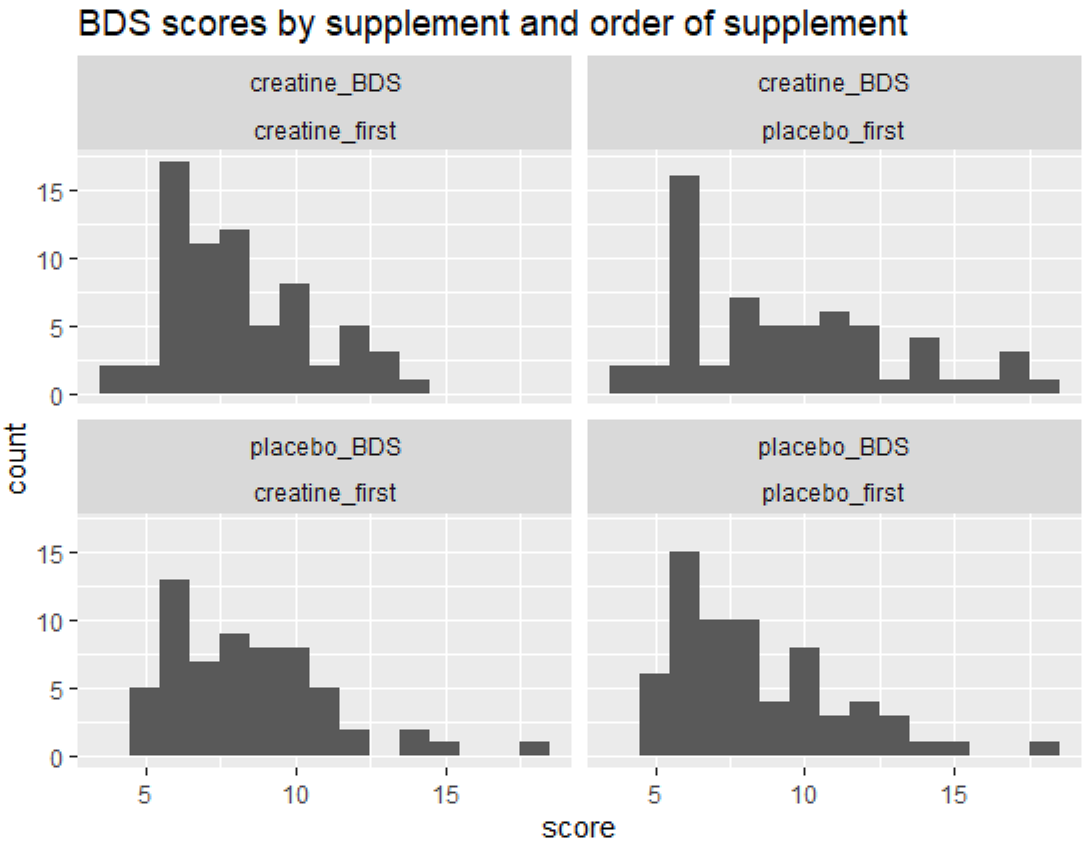

### Histograms of RAPM scores

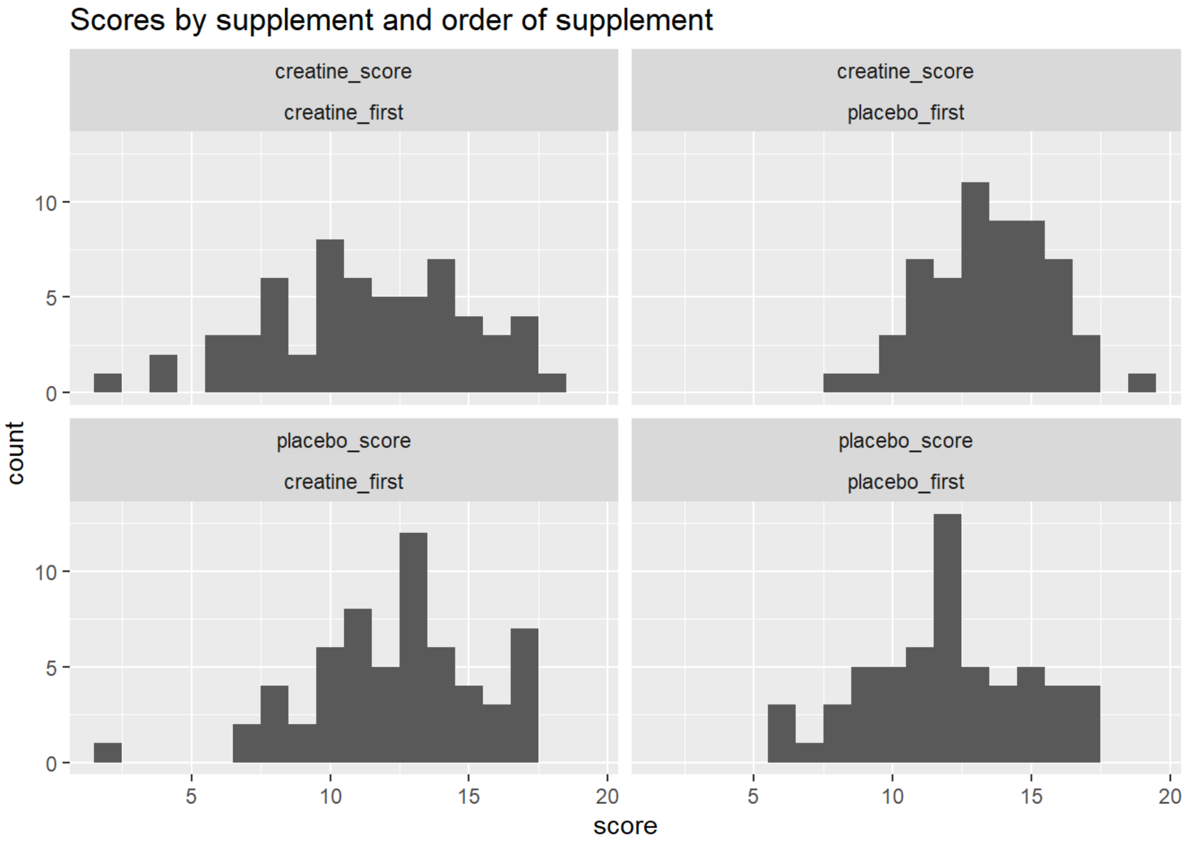


### Timeline graphs

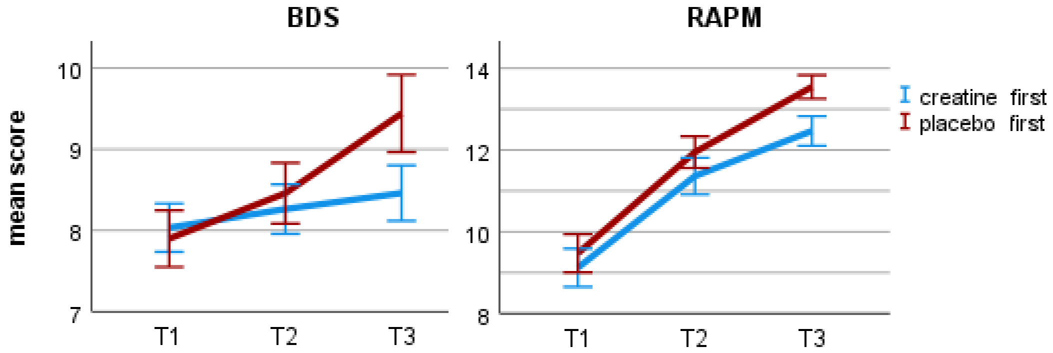


**Figure X.** a) Estimated marginal means for the Backward Digit Span (BDS) score at the first (baseline), second, and third testing. b) Estimated marginal means for Raven's Advanced Progressive Matrices (RAPM) score at the first (baseline), second, and third testing. Error bars represent 95% confidence intervals.

### Comparing baselines scores between supplement order groups

| Test_number | Test_name | p | higher baseline |
|---|---|---|---|
| Test01 | RAPM (abstract thought) | 0.594 | placebo_first |
| Test02 | BDS (working memory) | 0.77 | creatine_first |
| Test03 | blocktapping forward (working memory) | 0.077 | placebo_first |
| Test04 | blocktapping backward (working memory) | 0.718 | placebo_first |
| Test05 | BVMT-R (working memory) | 0.788 | creatine_first |
| Test14 | D2 | 0.008 | placebo_first |
| Test06 | FDS (working memory) | 0.607 | creatine_first |
| Test07 | Stroop - colors (reaction time) | 0.578 | creatine_first |
| Test08 | Stroop - colorletters (inhibition) | 0.88 | creatine_first |
| Test08minus07 | Stroop diff | 0.862 | placebo_first |
| Test09 | TMTA | 0.252 | creatine_first |
| Test10 | TMTB | 0.183 | creatine_first |
| Test11_1 | VLMT SumA1toA5 | 0.436 | placebo_first |
| Test11_2 | VLMT A5_minus_A6 (short term memory) | 0.716 | placebo_first |
| Test11_3 | VLMT A5_minus_A7 (long term memory) | 0.338 | placebo_first |
| Test12 | VLMT formula by Geffen 1990 (recognition memory) | 0.224 | placebo_first |
| Test13 | word fluency | 0.669 | placebo_first |

### Normality of BDS and RAPM

The BDS scores included two outliers and the RAPM scores included one outlier, defined as +/- 1.5 interquartile range from the third and first quarter, respectively. Skewness was above 1 (1.14 and 1.06) for two of the four factor combinations for the BDS scores and for no factor combination of the RAPM scores.

### Full results table BDS and RAPM

#### BDS

Threeway ANOVA

| Effect | Dfn | Dfd | F | p | pes | ges |
|---|---|---|---|---|---|---|
| diet | 1 | 117 | 0.182 | 0.671 | 0.002000 | 1.00e-03 |
| order_supplement | 1 | 117 | 1.329 | 0.251 | 0.011000 | 9.00e-03 |
| supplement | 1 | 117 | 3.412 | 0.067 | 0.028000 | 5.00e-03 |
| diet:order_supplement | 1 | 117 | 0.402 | 0.527 | 0.003000 | 3.00e-03 |
| diet:supplement | 1 | 117 | 0.060 | 0.808 | 0.000509 | 8.69e-05 |
| order_supplement:supplement | 1 | 117 | 7.586 | 0.007 | 0.061000 | 1.10e-02 |
| diet:order_supplement:supplement | 1 | 117 | 0.343 | 0.559 | 0.003000 | 4.99e-04 |

Twoway ANOVA

| N | N (c_first) | N (p_first) | **p (suppl.)** | p (suppl.*order) | p (order) | creatine score est. marg. mean, std. error | placebo score est. marg. mean, std. error |
|---|---|---|---|---|---|---|---|
| 121 | 61 | 60 | 0.064 | 0.007 | 0.246 | **8.85 (0.28)** | 8.44 (0.25) |

| Effect | Dfn | Dfd | F | p | pes | ges | SSn | SSd |
|---|---|---|---|---|---|---|---|---|
| (Intercept) | 1 | 119 | 1274 | 2.02e-65 | 0.915 | 0.899 | 18075.796 | 1687.956 |
| order_supplement | 1 | 119 | 1.358 | 0.246 | 0.011 | 0.009 | 19.267 | 1687.956 |
| supplement | 1 | 119 | 3.494 | 0.064 | 0.029 | 0.005 | 10.168 | 346.311 |
| order_supplement*supplement | 1 | 119 | 7.651 | 0.007 | 0.060 | 0.011 | 22.267 | 346.311 |

#### RAPM

Threeway ANOVA

| Effect | Dfn | Dfd | F | p | pes | ges |
|---|---|---|---|---|---|---|
| diet | 1 | 114 | 0.367 | 5.46e-01 | 0.003 | 0.00300 |
| order_supplement | 1 | 114 | 3.281 | 7.30e-02 | 0.028 | 0.02300 |
| supplement | 1 | 114 | 1.017 | 3.15e-01 | 0.009 | 0.00200 |
| diet:order_supplement | 1 | 114 | 0.530 | 4.68e-01 | 0.005 | 0.00400 |
| diet:supplement | 1 | 114 | 0.739 | 3.92e-01 | 0.006 | 0.00100 |
| order_supplement:supplement | 1 | 114 | 29.138 | 3.72e-07 | 0.204 | 0.04500 |
| diet:order_supplement:supplement | 1 | 114 | 0.268 | 6.06e-01 | 0.002 | 0.00043 |

Twoway ANOVA

| N | N (c_first) | N (p_first) | **p (suppl.)** | p (suppl.*order) | p (order) | creatine score est. marg. mean, std. error | placebo score est. marg. mean, std. error |
|---|---|---|---|---|---|---|---|
| 118 | 60 | 58 | 0.327 | < .001 | 0.075 | **12.39 (0.28)** | 12.16 (0.28) |

| Effect | Dfn | Dfd | F | p | pes | ges | SSn | SSd |
|---|---|---|---|---|---|---|---|---|
| (Intercept) | 1 | 116 | 2404 | 2.15e-79 | 0.954 | 0.944 | 35546.736 | 1714.997 |
| order_supplement | 1 | 116 | 3.225 | 7.50e-02 | 0.027 | 0.022 | 47.685 | 1714.997 |
| supplement | 1 | 116 | 0.968 | 3.27e-01 | 0.008 | 0.002 | 3.219 | 385.666 |
| order_supplement*supplement | 1 | 116 | 29.231 | 3.49e-07 | 0.201 | 0.044 | 97.185 | 385.666 |

### Post hoc tests of interaction in confirmatory analysis (learning effect)

Post-hoc t-tests of the interactions were conducted with an adjusted significance level of p = 0.05/4 = 0.013. For BDS, they find that participants in the creatine-first group do not improve significantly from timepoint 2 to timepoint 3 (p = 0.437) but participants in the placebo-first group do (p = 0.007), as would be expected if creatine had a beneficial effect. The difference in improvement is d = 0.36 - 0.1 = 0.26. For RAPM, both groups improve significantly from timepoint 2 to timepoint 3, most likely reflecting a learning effect. This improvement is more significant after creatine than placebo (p < .001 compared to p = 0.001) and the difference in improvement is d = 0.55 - 0.44 = 0.11.

| task | group | p | d | Creatine M (SD) | Placebo M (SD) |
|---|---|---|---|---|---|
| BDS | creatine-first | 0.437 | -0.1 | 8.26 (2.34) | **8.46 (2.67)** |
| BDS | placebo-first | 0.007 | 0.36 | **9.43 (3.63)** | 8.42 (2.87) |
| RAPM | creatine-first | 0.001 | -0.44 | 11.30 (3.65) | **12.35 (3.06)** |
| RAPM | placebo-first | < 0.001 | 0.55 | **13.48 (2.22)** | 11.87 (2.91) |

Table X.

### Cohen’s d - this study and Rae et al. (2003)

In order to compare our study more easily to the study of Rae et al. (2003), we estimated Cohen’s d for the two studies.

Cohen’s d = mean difference/SD of mean difference = mean difference/(SE of mean difference * sqrt(N))

For BDS in our study:
d = 0.41/(0.22*sqrt(121)) = 0.17

For RAPM in our study:
d = 0.23/(0.23*sqrt(118)) = 0.09

For BDS in Rae et al. (2003): 1 to 3 depending on calculation (see below)

For RAPM in Rae et al. (2003): 1 to 3 depending on calculation (see below)

Using this calculator and F and dfs:
- RAPM: Cohen’s f = 1.6, i.e. Cohen’s d = 3.2
- BDS: Cohen’s f = 1.5, i.e. Cohen’s d = 3

Using the same calculator and the raw M(SD) after-supplement scores (but unclear how balanced the order groups were and also Rae used the two baselines too, not just these scores):
- RAPM: Hedges g (basically the same as Cohen’s d) = 1 (sanity check: improvement in raw score = 4, sd = about 4, 4/4 = 1)
- BDS: Hedges g (basically the same as Cohen’s d) = 1 (sanity check: improvement in raw score = 1.5, sd = about 1.5, 1.5/1.5 = 1)

From Rae et al. (2003):

“Supplementation with oral creatine monohydrate significantly increased intelligence (as measured by RAPMs done under time pressure, figure 1a) compared with placebo (F(3 ,33) = 32.3, p , 0.0001; repeated-measures ANOVA). There was no significant effect of treatment order (F(1 ,33) = 1.62, p = 0.21), although there was a significant interaction with treatment order (F(3 ,99) = 6.7, p = 0.0004). The mean RAPMs raw score under placebo was 9.7 (s.d. = 3.8) items correct in 10 min versus 13.7 (s.d. = 4.1) items correct under the experimental treatment. Supplementation with oral creatine monohydrate (figure 1b) significantly affected performance on BDS (F(3 ,34) = 29.0, p < 0.0001), with no effect of order (F(3 ,102) = 0.98, p = 0.40). Mean BDS under the placebo was 7.05 items (s.d. = 1.19), compared with a mean of 8.5 items under creatine treatment (s.d. = 1.76).”

### Details on the Bayesian analysis

#### Models

Approach 1, point models: Approach 1 used point models for the null hypothesis and the alternative hypotheses. As the alternative hypothesis was not determined in advance and also to show to what extent the results are robust, we present the results for a range of alternative hypotheses. A point model means that we hypothesise a particular true effect size (0 for the null hypothesis, a number of small effect sizes for the alternative hypotheses). Even though a model postulates one particular effect size, the means expected to be sampled if this model was true would be normally distributed due to sampling error. So, entering a point model results in marginal model predictions that are normal and have the same standard deviation as the observed data.

Approach 2, half normal: Approach 2 compared a point null model against half normal distributions centred on zero and with the standard deviation set to half the maximum expected effect size. This distribution gives little weight to effect sizes that are larger than the maximum expected effect size and it gives more weight to an effect size the smaller it is.

For comparison, the Bayes factor calculations were also done with cauchy distributions.

#### Calculation 1 - point null and point alternative

The Bayes factor calculator used is here (https://bayesplay.colling.net.nz/) and the instructions on how to use it are here (http://www.lifesci.sussex.ac.uk/home/Zo).

We need the SE of the mean difference between creatine and placebo. To calculate it, I use SE = (mean difference)/t(supplement). To calculate t(supplement), I use t = sqrt(F(supplement)). F(supplement) is the F statistic for the supplement effect in the 2x2 ANOVA with supplement and supplement order.

Backward Digit Span

- F(supplement) = 3.494. So, t = sqrt(3.494) = 1.87.

- Mean difference (see estimated marginal mean of creatine and placebo score) = 8.85-8.44= 0.41.
- SE of mean difference = 0.41/1.87 = 0.22. Sanity check: SEs of creatine and placebo score are similar to SE of mean difference.

Raven's Advanced Progressive Matrices
- F(supplement) = 0.968. So, t = sqrt(0.968) = 0.984
- Mean difference (see estimated marginal mean of creatine and placebo score) = 12.39-12.16 = 0.23
- SE of mean difference = 0.23/0.984 = 0.23. Sanity check: Similar to SEs given for creatine and placebo score given by describe(my_data2).

Thus entering into the calculator:
- BDS
    - Normal data: mean = 0.41, SD = 0.22
    - Point null = 0
    - Point alternative = 0.1, 0.2, 0.4, 1.7
- RAPM
    - Normal data: mean = 0.23, SD = 0.23
    - Point null = 0
    - Point alternative = 0.1, 0.2, 0.4, 1.7

Results:

| Task | 0.1 | 0.2 | 0.4 | 1.7 |
|---|---|---|---|---|
| BDS | 2.1 | 3.6 | 5.7 | 2e-7 |
| RAPM | 1.4 | 1.6 | 1.3 | <2e-7 |

#### Calculation 2 - point null and half normal or Cauchy alternative

Data: same as in calculation 1 (normal, mean difference based on estimated means, SD = SE of mean difference)

Null: point 0. This translates to a normal distribution with a mean of 0 and an SE equal to the SE of my data, as can be seen in the advanced output of the calculator.

For the alternative models, set SD to half the size of the smallest unlikely effect, as recommended by Lincoln Colling in private correspondence.

Alternative, **small effect size**: half normal/cauchy with lower limit of 0. Smallest unlikely effect size in that world: mean difference in raw scores of 0.4-1 (d = 0.2 - 0.4), i.e. 0.4-1 more matrix solved or a 0.25-0.5 longer backward digit span. So, take 0.2-0.5 as SD/scale. Why d = 0.2-0.4 as the maximum? D = 0.2-0.4 would be 3-6 points in an IQ test. That is large for a longterm effect of a supplement and more noticeable than the effect is likely to be based on everyday experience.

- BDS, SD = 0.5
    - Half normal: BF(10) = 3.3
    - Half Cauchy: BF(10) = 2.3
- BDS, SD = 0.2
    - Half normal: BF(10) = 2.9
    - Half Cauchy: BF(10) = 2.6

- RAPM, SD = 0.5
    - Half normal: BF(10) = 1.0
    - Half Cauchy: BF(10) = 0.8
- RAPM, SD = 0.2
    - Half normal: BF(10) = 1.4
    - Half Cauchy: BF(10) = 1.1

Alternative, **Rae effect size**: half normal/Cauchy with lower limit of 0. Smallest unlikely effect size in that world: Twice that of Rae. So, take Rae's effect size (corresponding to a mean difference of 1.7 in the most conservative calculation) as SD/scale.
- BDS
    - Half normal: BF(10) = 1.4
    - Half Cauchy: BF(10) = 1.1
- RAPM
    - Half normal: BF(10) = 0.4
    - Half Cauchy: BF(10) = 0.3

### Bayesian analysis of Solis et al. (2014)

As mentioned in the discussion, we performed a Bayesian analysis of the data in Solis et al. (2014). Their result was: "vegetarians and omnivores had comparable brain total Cr content (5.999 (SD 0.811) v. 5.917 (SD 0.665) IU, respectively; P=0.77)".

Enter into calculator:
Normal data: mean = -0.082, SD = 0.197
Alternative models:
- point 0.3
- point 0.5
- half normal centred on 0 with SD = 0.3
- half normal centred on 0 with SD = 0.5

Null model: point 0

| Alternative model | Point 0.3 | Point 0.5 | half normal centred on 0 with SD = 0.3 | half normal centred on 0 with SD = 0.5 |
|---|---|---|---|---|
| BF(10) | 0.17 | 0.01 | 0.42 | 0.28 |

Calculations:

Data

- The difference in brain creatine content was 5.917-5.999 = -0.082.
- The participants were 14 vegetarians and 14 omnivores.
- The pooled SD was (0.665+0.811)/2 = 0.738.
- SE = SD/sqrt(N) = 0.738/sqrt(14) = 0.197

What are reasonable alternative hypotheses?

- The nine studies reviewed by Dolan et al. on the effect of supplementing creatine on brain creatine content found it increased by 5-10%.
- 5-10% for the brain creatine content measured by Solis et al. (2014) would be about 0.3 to 0.6 (6*0.05 to 6*0.1).

### Bayesian analysis of Solis et al. (2017)

The results were virtually the same as Solis et al. (2014) - the vegetarian group had a non-significantly *higher* brain creatine content. Same or very similar N, very similar p-value. The Bayes factors can only be very similar.

### Details on the analysis of diet

| Test_number | Test_name | p (suppl.* diet) | p (suppl.*diet * suppl-order) | p (suppl.) OMNI group | p (suppl.) VEG group |
|---|---|---|---|---|---|
| Test01 | RAPM (abstract thought) | 0.392 | 0.606 | 0.185 | 0.917 |
| Test02 | BDS (working memory) | 0.808 | 0.559 | 0.13 | 0.276 |
| Test03 | blocktapping forward (working memory) | 0.862 | 0.976 | 0.943 | 0.761 |
| Test04 | blocktapping backward (working memory) | 0.275 | 0.894 | 0.276 | 0.588 |
| Test05 | BVMT-R (working memory) | 0.806 | 0.636 | 0.557 | 0.792 |
| Test14 | D2 | 0.261 | 0.578 | 0.06 | 0.881 |
| Test06 | FDS (working memory) | 0.514 | 0.767 | 0.504 | 0.825 |
| Test07 | Stroop - colors (reaction time) | 0.601 | 0.974 | 0.557 | 0.856 |
| Test08 | Stroop - colorletters (inhibition) | 0.853 | 0.156 | 0.682 | 0.805 |
| Test08minus07 | Stroop diff | 0.923 | 0.174 | 0.889 | 0.705 |
| Test09 | TMTA | 0.008 | 0.223 | 0.003 | 0.471 |
| Test10 | TMTB | 0.206 | 0.584 | 0.427 | 0.322 |
| Test11_1 | VLMT SumA1toA5 | 0.495 | 0.916 | 0.713 | 0.553 |

| | | | | | |
|---|---|---|---|---|---|
| Test11_2 | VLMT A5_minus_A6 (short term memory) | 0.624 | 0.11 | 0.856 | 0.584 |
| Test11_3 | VLMT A5_minus_A7 (long term memory) | 0.71 | 0.71 | 0.389 | 0.586 |
| Test12 | VLMT formula by Geffen 1990 (recognition memory) | 0.799 | 0.492 | 0.672 | 0.944 |
| Test13 | word fluency | 0.235 | 0.391 | 0.989 | 0.13 |

#### Robust ANOVA with bootstrapping and 20% trim

| Test_number | Test_name | p (suppl.*diet) | p (suppl.) OMNI group | p (suppl.) VEG group |
|---|---|---|---|---|
| Test01 | RAPM (abstract thought) | 0.447 | 0.182 | 0.448 |
| Test02 | BDS (working memory) | 0.88 | 0.232 | 0.364 |

#### Bayesian analysis

Enter into calculator:

- Mean difference between veg group (creatine minus placebo) and omni group (creatine minus placebo), using estimated marginal means
- SE of mean difference

RAPM

- Mean difference: 0.0355-0.446 = -0.4105
    - Omni: (-0.965+1.857)/2 = 0.446
    - Veg: (-1.129+1.2)/2 = 0.0355
- SD of mean difference: (2.095+2.876+2.642+2.683)/4 = 2.57
- SE of mean difference: 2.57/sqrt(118) = 0.23

| diet <fctr> | order_supplement <fctr> | mean_cr_pl <dbl> | sd_cr_pl <dbl> | n <int> |
|---|---|---|---|---|
| O | creatine_first | -0.9655172 | 2.095621 | 29 |
| O | placebo_first | 1.8571429 | 2.876653 | 28 |
| V | creatine_first | -1.1290323 | 2.642498 | 31 |
| V | placebo_first | 1.2000000 | 2.683282 | 30 |

BDS

- Mean difference: 0.354-0.4625 = -0.1085
    - Omni: (-0.275+1.2)/2 = 0.4625
    - Veg: (-0.125+0.833)/2 = 0.354
- SD of mean difference: (1.943+2.618+2.012+2.995)/4 = 2.39
- SE of mean difference: 2.39/sqrt(121) = 0.22

| diet <fctr> | order_supplement <fctr> | mean_cr_pl <dbl> | sd_cr_pl <dbl> | n <int> |
|---|---|---|---|---|
| O | creatine_first | -0.2758621 | 1.943791 | 29 |
| O | placebo_first | 1.2000000 | 2.618238 | 30 |
| V | creatine_first | -0.1250000 | 2.012060 | 32 |
| V | placebo_first | 0.8333333 | 2.995207 | 30 |

Compare point null hypothesis to the same small effect sizes used for the Bayes factors of the confirmatory analysis in the manuscript and to the effect size of Benton and Donohoe (2011), who found that creatine was more beneficial for vegetarians than for omnivores.

Calculate the effect size of Benton and Donohoe (2011)
"However, after 4 d of consuming the creatine supplement, memory was better in vegetarians rather than in those who consumed meat ($F(1, 118) = 5.06$; $p < 0.03$)."

Using this calculator and F and dfs:

- memory: Cohen's f = 0.18, i.e. Cohen's d = 0.36
- D = 1 corresponds to a difference in raw scores of 2.5 in our study.
- I.e. d = 0.36 corresponds to about 2.5/3 = 0.8 difference in raw scores in our study.

| | **Approach 1: point models** | | | | **Approach 2: half normal** | | |
|---|---|---|---|---|---|---|---|
| | Small effects | | | BD-sized | Small effects | | Max. = 2xBD-size |
| **Task** | **0.1** | **0.2** | **0.4** | **0.8** | **max. 0.4** | **max. 1** | **max. 1.6** |
| BDS | 0.72 | 0.42 | 0.08 | < 2e-7 | 0.58 | 0.29 | 0.19 |
| RAPM | 0.42 | 0.15 | 0.01 | < 2e-7 | 0.36 | 0.16 | 0.10 |

### Details on the analysis of exploratory cognitive tasks

There was no indication that creatine improved performance in our additional exploratory cognitive tasks.

| Test_name | N | N (c_first) | N (p_first) | **p (suppl.)** | p (suppl.*order) | p (order) | creatine score est. marg. mean, std. error | placebo score est. marg. mean, std. error |
|---|---|---|---|---|---|---|---|---|
| blocktapping forward (working memory) | 71 | 37 | 34 | 0.779 | 0.779 | 0.591 | **10.61 (0.26)** | 10.54 (0.24) |
| blocktapping backward (working memory) | 70 | 37 | 33 | 0.83 | 0.525 | 0.513 | 9.62 (0.31) | **9.69 (0.24)** |
| BVMT-R (working memory) | 119 | 61 | 58 | 0.543 | 0.997 | 0.859 | **31.42 (0.41)** | 31.19 (0.45) |
| D2 | 104 | 51 | 53 | 0.394 | < .001 | 0.363 | 226.27 (4.19) | **227.91 (4.23)** |
| FDS (working memory) | 117 | 60 | 57 | 0.714 | 0.133 | 0.796 | 9.28 (0.22) | **9.34 (0.22)** |
| Stroop - colors (reaction time) | 118 | 60 | 58 | 0.813 | 0.039 | 0.885 | 44.45 (1.20) | **44.70 (1.22)** |
| Stroop - colorletters (inhibition) | 119 | 60 | 59 | 0.626 | 0.05 | 0.971 | 67.59 (1.38) | **67.12 (1.28)** |
| Stroop diff | 118 | 60 | 58 | 0.736 | 0.415 | 0.883 | 23.00 (0.97) | **22.66 (0.94)** |
| TMTA | 123 | 62 | 61 | 0.129 | 0.106 | 0.393 | 22.54 (0.63) | **21.76 (0.65)** |
| TMTB | 122 | 62 | 60 | 0.855 | 0.004 | 0.074 | **53.12 (1.98)** | 53.41 (2.01) |
| VLMT SumA1toA5 | 119 | 60 | 59 | 0.87 | 0.49 | 0.289 | **63.20 (0.73)** | 63.11 (0.76) |
| VLMT A5_minus_A6 (short term memory) | 119 | 60 | 59 | 0.847 | 0.454 | 0.069 | **1.11 (0.15)** | 1.14 (0.17) |
| VLMT A5_minus_A7 (long term memory) | 118 | 60 | 58 | 0.301 | 0.389 | 0.478 | 1.38 (0.17) | **1.18 (0.17)** |
| VLMT formula by Geffen 1990 (recognition memory) | 117 | 59 | 58 | 0.722 | 0.017 | 0.159 | **0.86 (0.1)** | 0.85 (0.1) |
| word fluency | 122 | 60 | 62 | 0.227 | 0.928 | 0.435 | **25.47 (0.60)** | 24.84 (0.61) |

| | BWTRIM (20% trimmed ANOVA) | winsorised 5% | winsorised 20% | **SPPB (bootstrapped and 20% trimmed ANOVA)** |
|---|---|---|---|---|
| Test_name | p (suppl.) | p (suppl.) | p (suppl.) | **p (suppl.)** |
| blocktapping forward (working memory) | 0.564 | 0.865 | 0.678 | 0.826 |
| blocktapping backward (working memory) | 0.87 | 0.482 | 0.482 | 0.59 |
| BVMT-R (working memory) | 0.809 | 0.746 | 0.112 | 0.67 |
| D2 | 0.382 | 0.446 | 0.291 | 0.62 |
| FDS (working memory) | 0.795 | 0.838 | 0.721 | 0.52 |
| Stroop - colors (reaction time) | 0.184 | 0.432 | 0.054 | 0.568 |
| Stroop - colorletters (inhibition) | 0.877 | 0.861 | 0.547 | 0.856 |
| TMTA | 0.040 | 0.068 | 0.021 | 0.224 |
| TMTB | 0.622 | 0.745 | 0.567 | 0.560 |
| VLMT SumA1toA5 | 0.744 | 0.996 | 0.883 | 0.996 |
| VLMT A6 (short term memory) | 0.622 | 0.854 | 0.323 | 0.806 |
| VLMT A7 (long term memory) | 0.346 | 0.462 | 0.133 | 0.540 |
| VLMT formula by Geffen 1990 (recognition memory) | 0.327 | 0.398 | 0.635 | 0.348 |
| word fluency | 0.631 | 0.272 | 0.119 | 0.692 |

### Second supplementation and high-baseline participants

To investigate the effect of the first supplementation, we conducted a mixed ANOVA with time (baseline and testing after first supplementation) as the within-subjects variable, first supplement as the between-subjects variable and test score as the dependent variable. For the second supplementation, the same ANOVA was conducted except with time (baseline and testing after second supplementation) as the within-subjects variable and second supplement as the between-subjects variable. We performed these ANOVAs with the assumption of normality as well as a robust version.

To investigate the effect of baseline performance, we split participants at the median score (for each cognitive task separately) into low and high baseline participants and conducted separate ANOVAs for each group.

For both cognitive tasks, the effect of creatine was more positive after the second supplementation than the first supplementation. This pattern was more pronounced for BDS than for RAPM (Table X). For BDS but not RAPM, high baseline participants showed a robust creatine effect while low baseline participants did not. These exploratory results suggest that creatine might have a greater effect when performance at or familiarity with a task is high. Of course, this analysis was not preregistered and the result may not reproduce.

| **Task** | **p (creatine effect)** | | | | | | | | | | | |
|---|---|---|---|---|---|---|---|---|---|---|---|---|
| | Best | normal | robust | Best | normal | robust | Best | normal | robust | Best | normal | robust |
| | **First supplementation** | | | **Second supplementation** | | | **Low baseline participants** | | | **High baseline participants** | | |
| RAPM | pl | 0.649 | 0.476 | crea | 0.221 | 0.19 | crea | 0.837 | 0.872 | crea | 0.25 | 0.966 |
| BDS | pl | 0.395 | 0.846 | crea | 0.013 | 0.246 | pl | 0.971 | 0.862 | crea | 0.018 | 0.052 |

**Table X.** Creatine effect p-values (two-tailed) for different ANOVAs, looking at the first and the second supplementation separately and at low- and high-baseline participants separately. Better score given as pl = placebo and crea = creatine. Only complete participants are included. The robust ANOVA uses bootstrap and 20% trim. For the first and second supplementation ANOVAs, the creatine effect is the supplement*time interaction. Time includes the baseline and the score after the first or second supplementation, respectively. For the ANOVAs split by baseline, the creatine effect is the supplement main effect.

### Exploratory tasks: Low vs high baseline participants

| Test_name | p (suppl.) LOW baseline group | p (suppl.) HIGH baseline group | same direction |
|---|---|---|---|
| blocktapping forward (working memory) | 0.822 | 0.852 | yes |
| blocktapping backward (working memory) | 0.176 | 0.244 | no |
| BVMT-R (working memory) | 0.931 | 0.116 | no |
| D2 | 0.633 | 0.336 | yes |
| FDS (working memory) | 0.124 | 0.875 | no |
| Stroop - colors (reaction time) | 0.695 | 0.978 | yes |
| Stroop - colorletters (inhibition) | 0.735 | 0.469 | no |
| Stroop diff | 0.847 | 0.602 | no |
| TMTA | 0.062 | 0.972 | NA |
| TMTB | 0.282 | 0.488 | no |
| VLMT SumA1toA5 | 0.867 | 0.554 | no |
| VLMT A5_minus_A6 (short term memory) | 0.346 | 0.709 | no |
| VLMT A5_minus_A7 (long term memory) | 0.8 | 0.113 | no |
| VLMT formula by Geffen 1990 (recognition memory) | 0.87 | 0.493 | no |
| word fluency | 0.404 | 0.031 | no |

### Demographic and context variables

Participants filled out a questionnaire on their daily condition (including sleep, food, caffeine, and infections) before each testing. Before the baseline testing, they filled out the Edinburgh Handedness Inventory (Oldfield, 1971) and performed a test of crystallised intelligence called “Mehrfach-Wahl-Wortschatztest (MWT-B)” (Lehrl, 2005).

There was no indication that women benefited more from creatine than did men. There was not enough variation in adherence nor in sleep to justify analysing an effect of these variables. There was no indication that creatine affected different age groups differently, but this could very well be because we did not have enough older participants to study this question well (median = 28 years). We did not investigate any other demographic or context variables.

### Details on reasons for correct guesses of supplement order

The only participants we asked were the 33 second round participants

- 11 of those stated solubility among their reasons for their guess and of those, 9(!) guessed correctly
- 4 people mentioned negative side effects as a reason and all of them guessed correctly. One of them had also mentioned solubility as a reason.
- 9 people mentioned positive side effects among their reasons and of those, 6 guessed correctly. 3 of them had also mentioned solubility (and guessed correctly)

--> Solubility and negative side effects seem to lead to correct guesses, while just positive effects don't. But solubility was mentioned almost three times more often as a reason than negative effects. However, when we break this down further we can see that two participants mentioned as positive effects a clear increase in muscle and they guessed correctly. These were two participants who exercised hard and almost exceeded our exercise limit. I don’t think their correct guess was a coincidence.
